# Influence of clinical characteristics and anti-cancer therapy on outcomes from SARS-CoV-2 infection: a systematic review and meta-analysis of 5,678 cancer patients

**DOI:** 10.1101/2020.12.15.20248195

**Authors:** Ik Shin Chin, Sara Galavotti, Kay Por Yip, Helen Curley, Roland Arnold, Archana Sharma-Oates, Laura Chegwidden, Siang Ing Lee, Lennard YW Lee, David J. Pinato, Gino M. Dettorre, Claire Palles

**Author notes:** **Corresponding Author:** Ik Shin Chin, Institute of Cancer and Genomic Sciences, University of Birmingham, Edgbaston, Birmingham B15 2TT, UK, 0121 414 3344.

## Abstract

**Background:** The COVID-19 pandemic started a healthcare crisis and heavily impacted cancer services.

**Methods:** Data from cohort studies of COVID-19 cancer patients published up until October 23rd 2020 from PubMed, PubMed Central, medRxiv and Google Scholar were reviewed. Meta-analyses using the random effects model was performed to assess the risk of death in cancer patients with COVID-19.

**Results:** Our meta-analyses including up to 5,678 patients from 13 studies showed that the following were all statistically significant risk factors for death following SARS-CoV-2 infection in cancer patients: age of 65 and above, presence of co-morbidities, cardiovascular disease, chronic lung disease, diabetes and hypertension. There was no evidence that patients who had received cancer treatment within 60 days of their COVID-19 diagnosis were at a higher risk of death, including patients who had recent chemotherapy.

**Conclusions:** Cancer patients are susceptible to severe COVID-19, especially older patients and patients with co-morbidities who will require close monitoring. Our findings support the continued administration of anti-cancer therapy during the pandemic. The analysis of chemotherapy was powered at 70% to detect an effect size of 1.2 but all other anti-cancer treatments had lower power. Further studies are required to better estimate their impact on the outcome of cancer patients.

## Introduction

SARS-CoV-2 which has led to the COVID-19 pandemic has resulted in over 65 million COVID-19 cases and 1.5 million deaths globally as of 6 December 2020 [1][2][3]. Cancer patients are considered to be at high risk from COVID-19 compared to the general population due to underlying immunosuppression from their malignancy and anti-cancer treatment. A large cohort study of over 20,000 hospitalised COVID-19 patients in the UK identified 1,743 (10%) patients with a cancer diagnosis, which was significantly associated with hospital mortality (adjusted HR 1.13, 95% CI 1.02-1.24, p=0.017) [4]. Cancer services was intentionally restricted in many countries at the beginning of the pandemic as a precaution. Since then, an estimated 20% increase in mortality in patients with new cancer diagnoses has been attributed to delays in diagnosis and treatment as a consequence of COVID-19 [5]. A recent survey conducted by Cancer Research UK found that 1 in 3 cancer patients reported that their treatment had been affected by the pandemic [6]. Balancing the benefits and risks of each patient’s cancer treatment by taking into account factors such as treatment survival and response benefit, co-morbidities, prognosis, as well as the risks of immunosuppression are important considerations but often challenging [7]. We aimed to investigate the impact of cancer patients’ characteristics, cancer type and recent treatment history on the risk of death in cancer patients following SARS-CoV-2 infection.

## Methods

### Literature review strategy and inclusion criteria

The review was conducted according to the PRISMA guidelines (Figure 1, Supplementary Table 1) [8]. PubMed and PubMed Central (PMC) were searched for articles published in the last year up to 23^rd^ October 2020 using the following keywords: coronavirus, COVID-19, SARS-CoV-2 AND cancer, malignancy and oncology. The same keywords were also used to identify relevant articles using medRxiv and Google Scholar. All articles available in English were screened. Studies reporting on characteristics, treatment history and outcome of cancer patients diagnosed with COVID-19 from real-time polymerase chain reaction (RT-PCR) testing or clinical features were included. Minimum inclusion criteria were: reporting on 10 or more COVID-19 positive cancer patients and provision of data on sex and age. For studies that had potentially overlapping participants based on the hospital recruitment sites and authors’ institutions listed, the study with the largest number of participants was included. Studies that contained abstracts only or analysed specific cancer types only or paediatric cancers were excluded.

**Figure 1.**
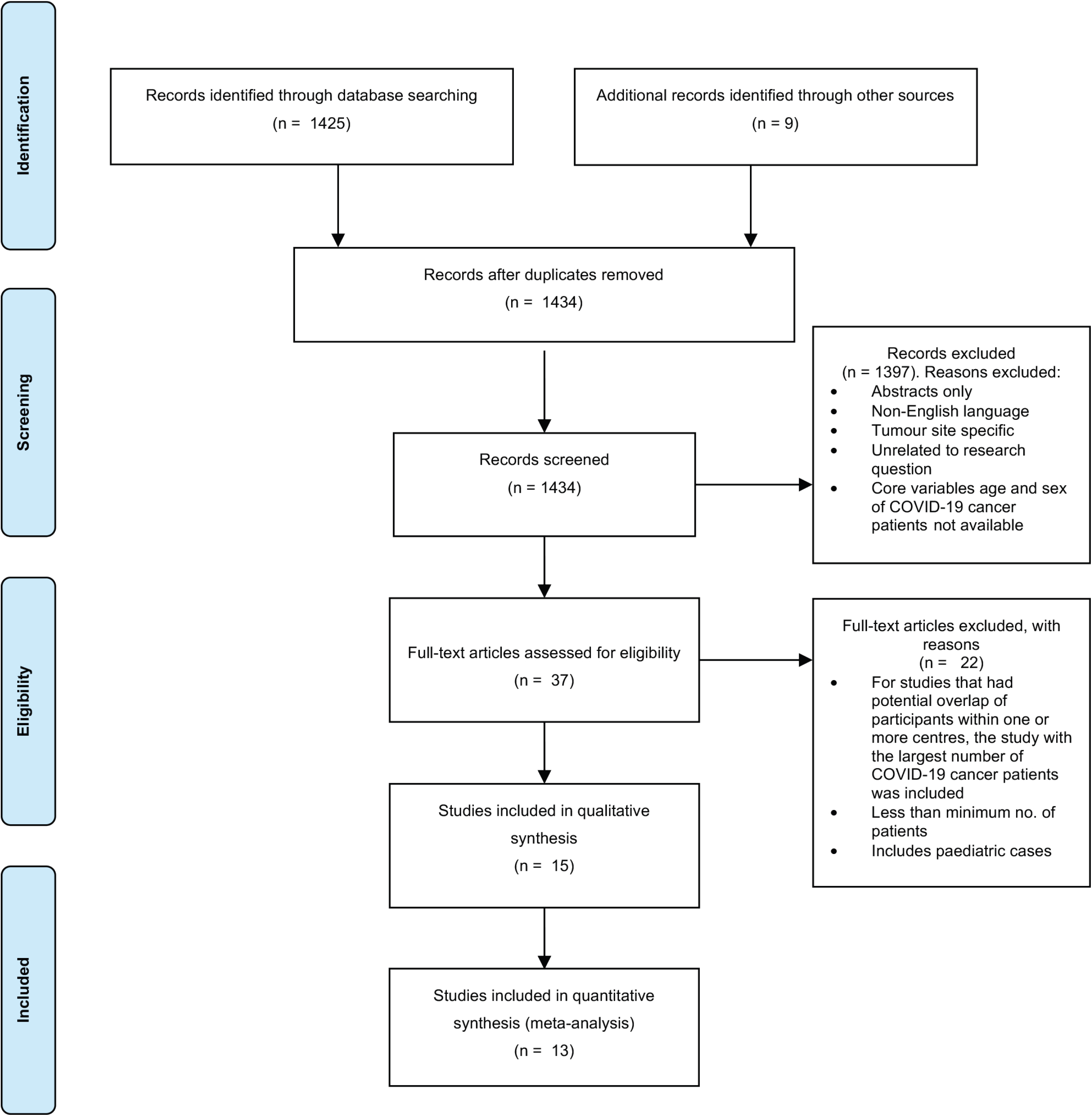
PRISMA (Preferred Reporting Items for Systematic Reviews and Meta-Analyses) flowchart showing how the included studies were identified.

### Data extraction process and statistical analysis

Two independent reviewers were allocated for each study. Counts of the number of patients that died and the number that recovered following SARS-CoV-2 infection, stratified by the following variables were extracted from each of the source data papers or requested from the corresponding author: sex, age, co-morbidities, Eastern Cooperative Oncology Group performance status (ECOG PS), smoking status, cancer type and stage and anti-cancer therapy. Count data was extracted for all-cause mortality and deaths caused by COVID-19 where available. Two of the registry studies identified (UKCCMP and OnCovid) had two publications each. Data from the more recent publication was extracted where the same variables had been reported with updated data [9][10][11][12]. Raw, previously unpublished data was provided by the OnCovid team to allow formal meta-analyses of key variables such as cancer treatment types. In all meta-analyses, studies were only included if they had provided counts data on patients with each outcome and variable under investigation to allow fair comparison across studies.

Meta-analyses of binary outcome data with ‘events’ corresponding to the number of deaths were performed with the ‘R’ statistical packages *meta*(*v4*.*15-1*) and *metafor*(*v2*.*4-0*) [13][14]. Random effects model was used as the studies in the meta-analyses originated from multiple different populations. The Mantel-Haenszel method was selected for data pooling using risk ratios (RR) as the summary measure. Between-study heterogeneity was tested using the Sidik-Jonkman estimator. Publication bias was assessed using funnel plots and funnel plot asymmetry was quantified with Egger’s test if necessary. Power calculations for each analysis was performed using *epiR*(*v1*.*0-15*) package from ‘R’.

## Results

### Description of the included studies

15 papers published describing a total of 6,711 cancer patients with COVID-19 met the inclusion criteria. The number of COVID-19 cancer patients included in each study ranged from 25 to 1289 (Table 1). The search and screening strategies used to identify each study is shown in Figure 1 and the study characteristics and variables measured by each are summarised in Table 1. Ten studies were national or international multi-centre studies involving sites from the United Kingdom (UK), United States (US), Italy, Spain, Belgium, Germany, Netherlands, China, Canada and France. The remaining five studies were single-centre studies originating from India, Italy, US, UK and Brazil. Studies focusing on a specific cancer type were excluded but two studies focusing on patients with solid cancers were included [15][16]. Cancer patients were diagnosed with COVID-19 based on RT-PCR testing alone in 10/13 (77%) of the studies (excluding two earlier publications from UKCCMP and OnCovid). The remaining studies used clinical or radiological approaches to confirm the diagnosis. All studies reported on the number of deaths in COVID-19 cancer patients. Mortality rates reported from the studies reviewed ranged from 13% to 60.4%. However, not all studies reported COVID-19 related deaths only (Table 1).

**Table 1.**
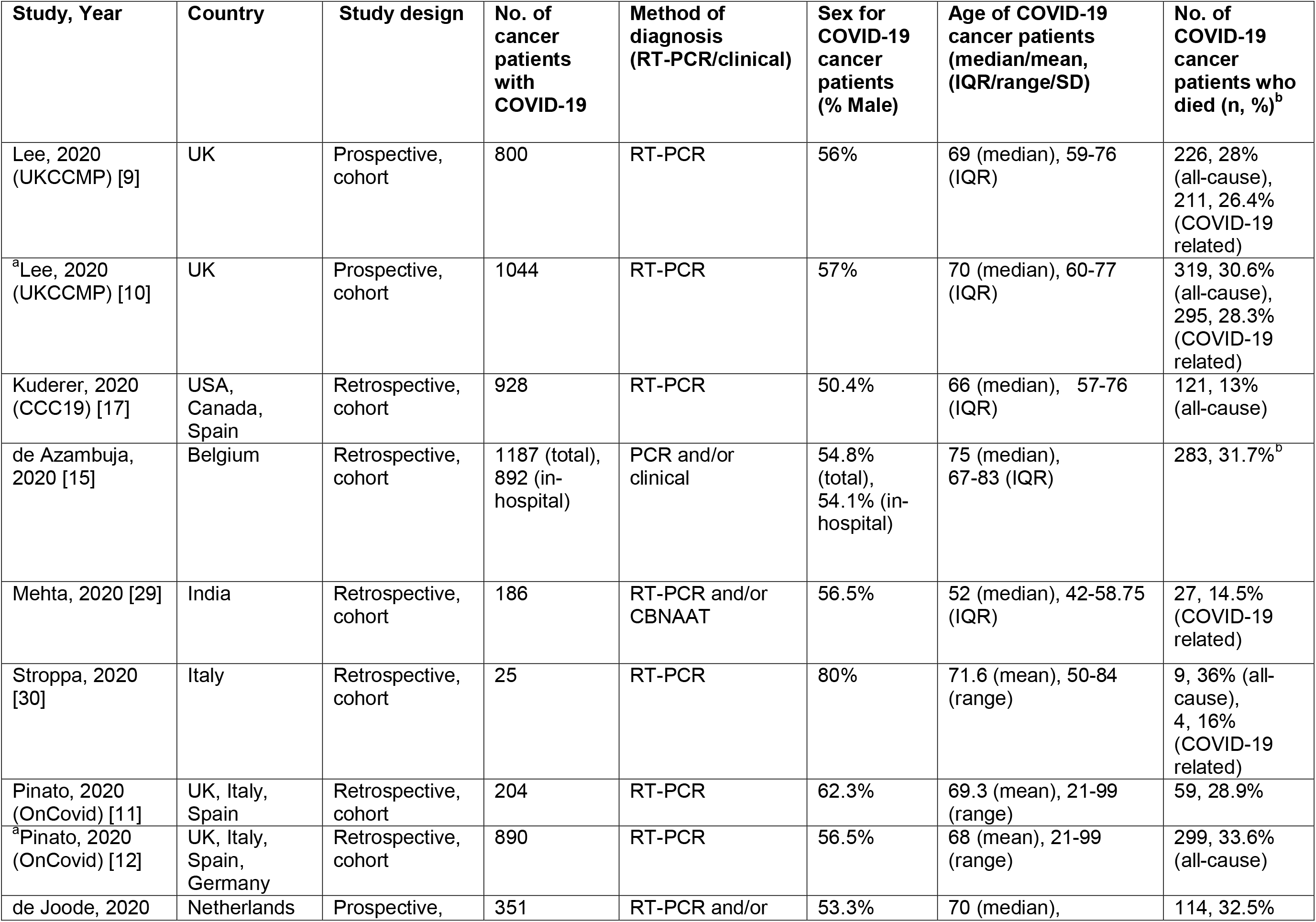
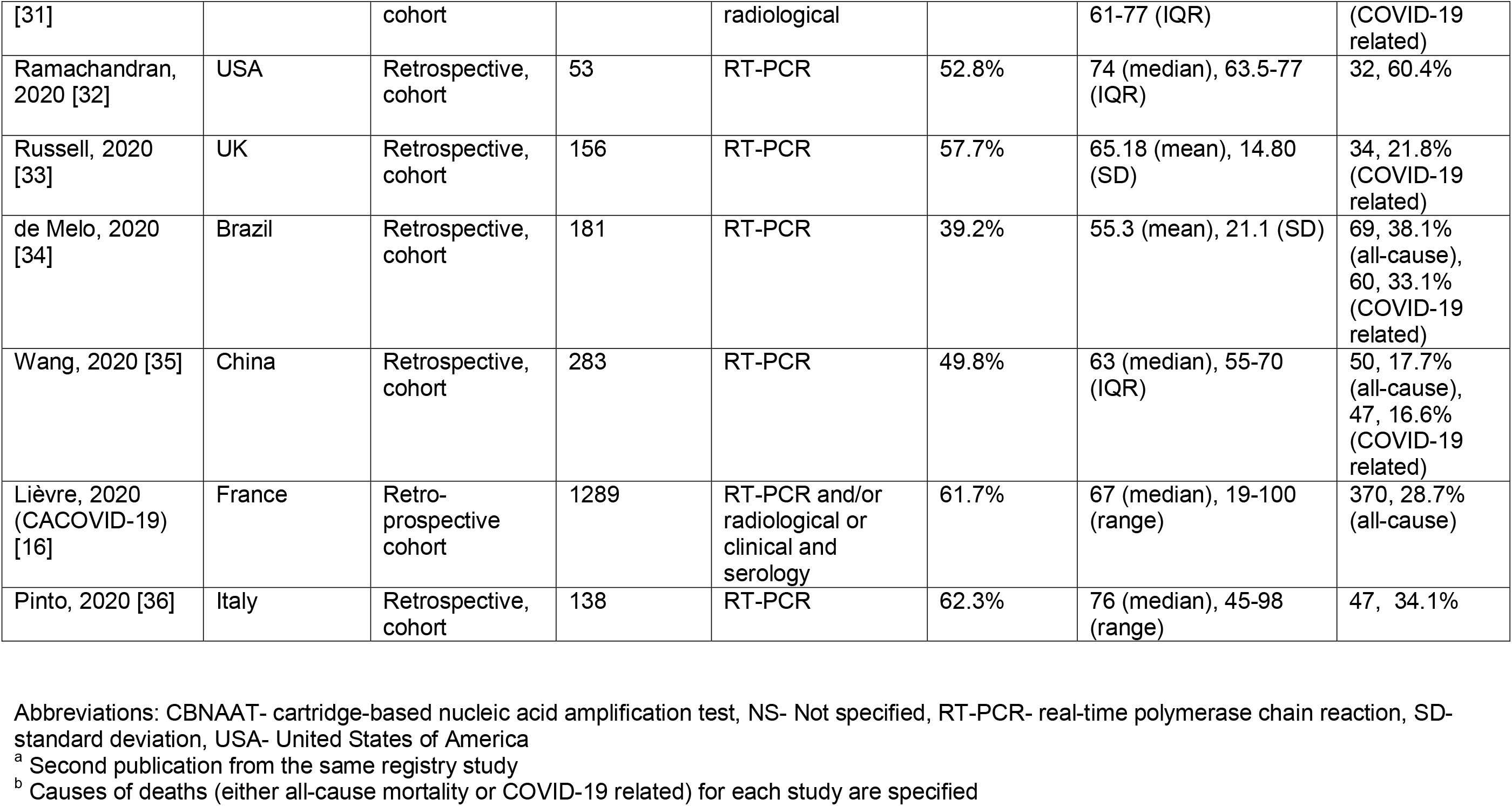
Characteristics of the included studies and variables of sex, age and mortality rates of cancer patients with COVID-19.

### Characteristics of COVID-19 positive cancer patients

#### Age and sex

Age and sex of COVID-19 cancer patients were reported in all the studies reviewed. The pooled proportion of male COVID-19 positive cancer patients is 0.55 (95% confidence interval (CI) 0.51-0.60) with significant heterogeneity (*I*^*2*^ 81%, p<0.01) (Supplementary Fig. 1). A meta-analysis from 10 studies showed that male cancer patients had a higher risk of death compared to female cancer patients although this was not statistically significant (Risk Ratio(RR) 1.25, 95% CI 0.91-1.72, p-value=0.15) (Supplementary Fig. 2). All studies reported either the median or mean age of COVID-19 positive cancer patients analysed, which ranged between age 52 to 76 and 55.3 to 71.6 respectively. Four studies showed a statistically significant higher risk of death in patients who are of age 65 or older (RR 2.27, 95% CI 1.12-4.62, p=0.04), but with significant heterogeneity between studies (*I*^*2*^ 84.1%, p=0.0003) (Figure 2).

**Figure 2.**
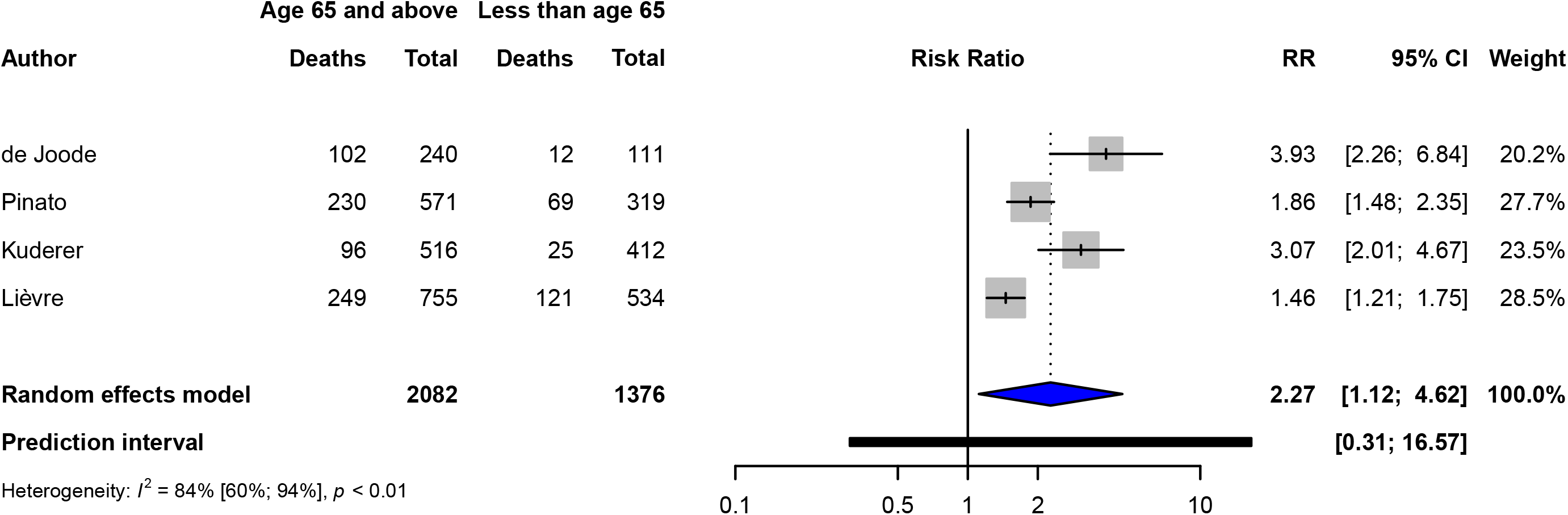
Forest plot of contribution of age to risk of death in COVID-19 cancer patients when comparing those aged 65 or older to patients less than age 65. Abbreviations: CI-Confidence interval, RR-Risk ratio

#### Performance status and smoking status

The outcomes of cancer patients with COVID-19 according to ECOG PS were reported in three studies. There was a non-significant higher risk of death in cancer patients with a PS of 3 or greater (RR 2.32, 95% CI 0.64-8.38, p=0.11) with considerable heterogeneity (*I*^*2*^ 73%, p=0.02) (Supplementary Fig. 3). Four studies reported on outcomes for patients with and without a smoking history. On analysis, there was a non-significant association between smoking history and an increased risk of death (RR 1.37, 95% CI 0.76-2.49, p=0.19) (Supplementary Fig. 4).

**Figure 3.**
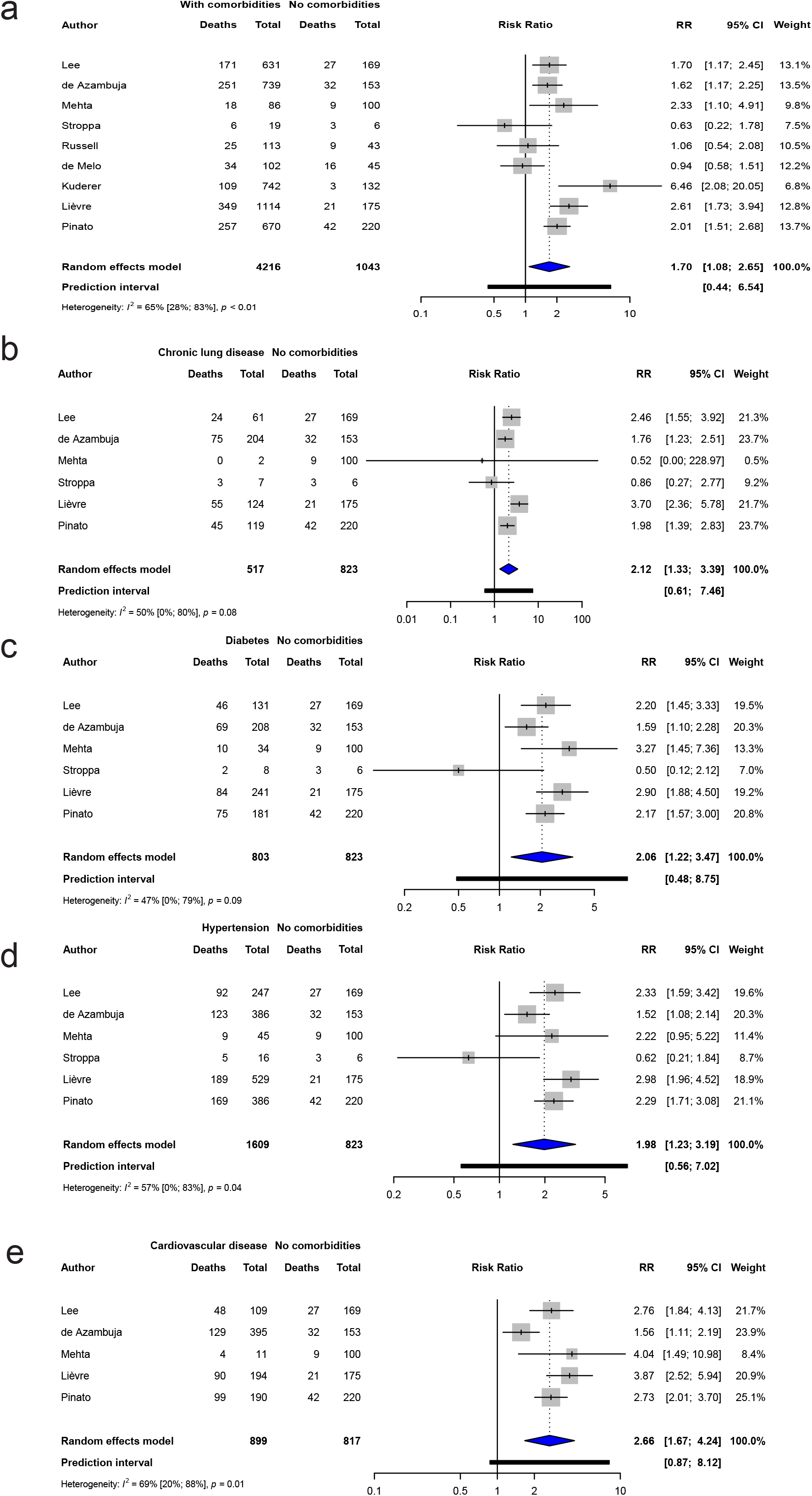
Forest plot of the contribution of co-morbidities to risk of death in COVID-19 cancer patients. a) Those with one or more co-morbidity were compared to those with no co-morbidities. b – e) Further analysis was performed to investigate the contribution of common co-morbidities including chronic lung disease, diabetes, hypertension and cardiovascular disease. Abbreviations: CI-Confidence interval, RR-Risk ratio

**Figure 4.**
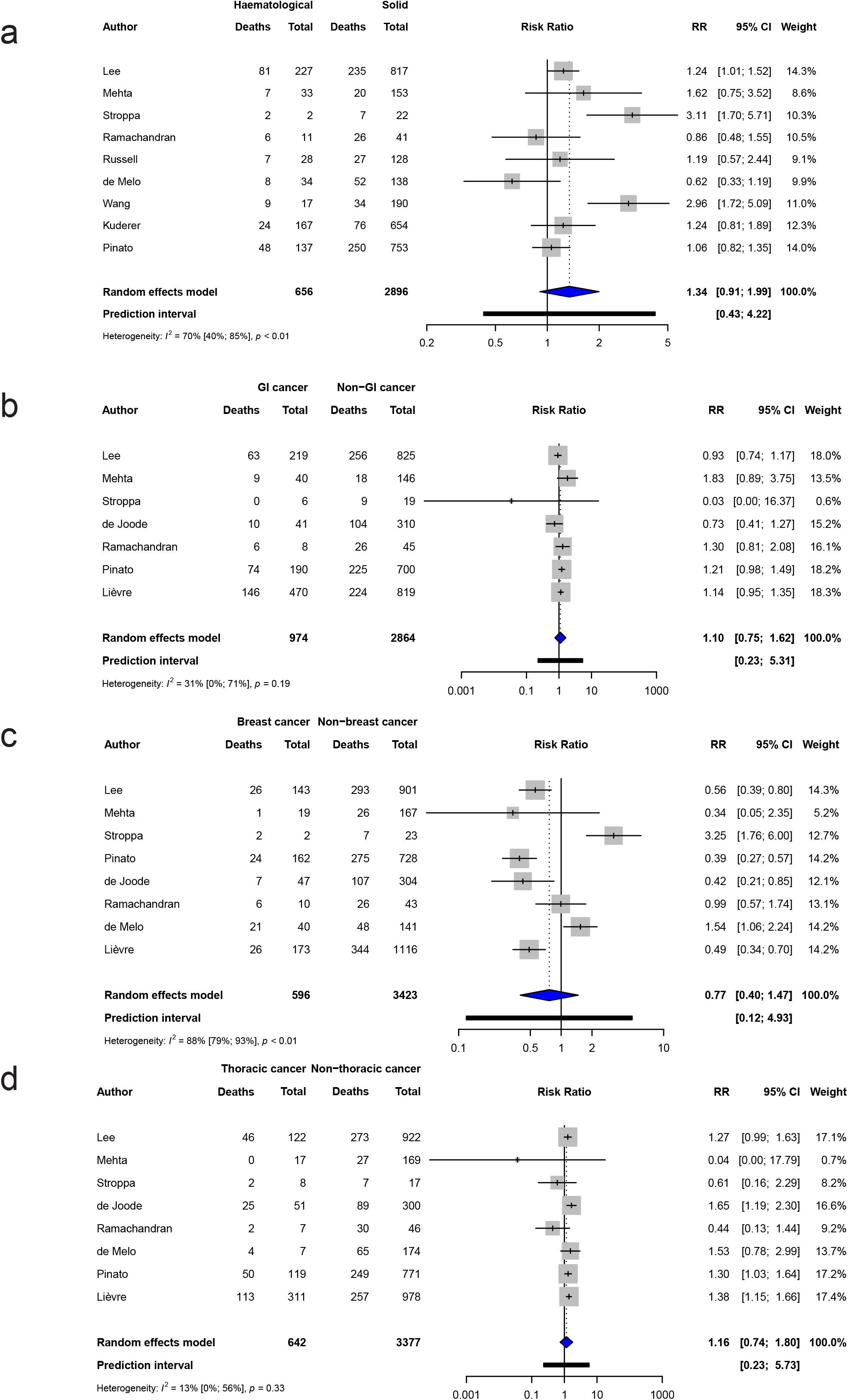
Forest plot of risk of death in COVID-19 cancer patients when considering different cancer types. a) Cancer patients with solid cancer compared to haematological cancers. b-d) Gastrointestinal, breast and thoracic cancer patients were compared to the rest of the patients in each study. Abbreviations: CI-Confidence interval, GI-Gastrointestinal, RR-Risk ratio

#### Presence and type of comorbidities

11 studies reported on the proportion of cancer patients with co-morbidities. Between 46% and 86% of the cancer patients in these studies had one or more co-morbidities (Supplementary Table 2). Meta-analysis from these studies showed a significant higher risk of death in patients with co-morbidities (RR 1.70, 95% CI 1.09-2.65, p=0.03) with significant inter-study heterogeneity (*I*^*2*^ 64.9%, p=0.004) (Figure 3a). Six studies provided summary data on specific co-morbidities. Chronic lung disease, diabetes and hypertension were all significantly associated with higher risk of death (RR 2.12, 95% CI 1.33-3.39, p=0.009, RR 2.06, 95% CI 1.22-3.47, p=0.02 and RR 1.98, 95% CI 1.23-3.19, p=0.01 respectively) with heterogeneity only significant for hypertension scores (*I*^*2*^ 57%, p=0.04) (Figure 3b-d). A meta-analysis of five studies showed that cardiovascular disease was significantly associated with an increased risk of death (RR 2.66, 95% CI 1.67-4.24, p=0.004) with also significant heterogeneity (*I*^*2*^ 68.7%, p=0.01) (Figure 3e).

#### Cancer stage and cancer type

12 studies described the proportion of COVID-19 cancer patients with metastatic cancer. This ranged from 24.5% to 58.8% (Supplementary Table 2). A meta-analysis of five studies showed that metastatic cancer was associated with a higher risk of death, but did not meet statistical significance (RR 1.53, 95% CI 0.98-2.37, p=0.06) (Supplementary Fig. 5). However, three of the studies reported on all-cause mortality, while two studies reported COVID-19 related deaths. Therefore we were unable, in the former studies, to exclude the possibility that excess death risks could be attributed to cancer related deaths. 12 studies reported on the cancer types in their patient cohort. A meta-analysis of nine studies found a non-significant increased risk of death in haematological cancer patients (RR 1.34, 95% CI 0.91-1.99, p=0.12) (Figure 4a). Four studies reported haematological cancers as the most frequent cancer type among patients with COVID-19. Meta-analysis of seven studies showed that gastrointestinal cancers were not significantly associated with an increased risk of death (RR 1.10, 95% CI 0.75-1.62, p=0.56) (Figure 4b). Eight studies also showed no significant association between breast cancer and risk of death (RR 0.77, 95% CI 0.40-1.47, p=0.36) but there was significant heterogeneity (*I*^*2*^ 88%, P<0.0001) (Figure 4c). Similarly, eight studies showed no significant association between death and thoracic cancers (RR 1.16, 95% CI 0.75-1.80, p=0.46) (Figure 4d). As the study by Lièvre only analysed solid cancers, we repeated the meta-analyses of gastrointestinal, breast and lung cancer without this study. The results were similar but remained non-significant (Supplementary Table 3).

**Figure 5.**
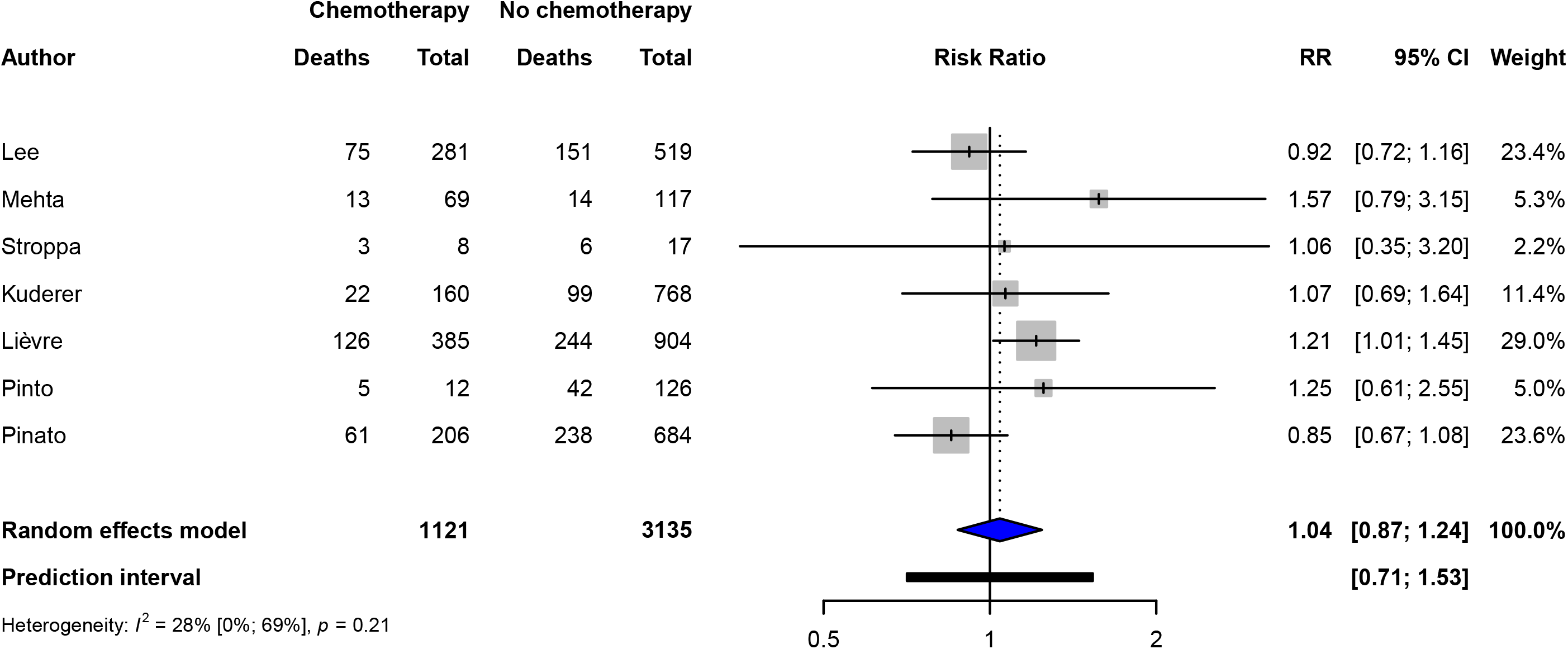
Forest plot of risk of death in COVID-19 cancer patients who had recent chemotherapy. Recent was defined as ongoing treatment at time of COVID-19 diagnosis or within 60 days of their diagnosis. Abbreviations: CI-Confidence interval, RR-Risk ratio

### Impact of anti-cancer treatment

12 studies provided information on cancer treatment history prior to COVID-19 infection. 10% to 65% of cancer patients received recent anti-cancer therapy prior to admission (Supplementary Table 2). The definition of recent treatment varied significantly between studies. A meta-analysis of six studies defining recent anti-cancer treatment as either receiving active treatment or within 60 days prior to infection showed a borderline significant reduction in the risk of death in those receiving anti-cancer treatment (RR 0.83, 95% CI 0.68-1.02, p=0.07) (Supplementary Fig. 6a). 11/13 studies had data on the types of anti-cancer therapy received. Chemotherapy was the most commonly administered in 9% to 37% of patients. A meta-analysis of seven studies demonstrated no significant association between recent chemotherapy and death (RR 1.04, 95% CI 0.88-1.24, p=0.59) (Figure 5). In addition, six studies showed no significant association between recent immunotherapy and risk of death (RR 0.94, 95% CI 0.63-1.43, p=0.73) and five studies similarly showed no association between recent targeted therapy and risk of death (RR 0.99, 95% CI 0.50-1.95, p=0.96) (Supplementary Fig. 6b & c). Recent radiotherapy and hormonal therapy were also not associated with a higher risk of death (RR 0.83, 95% CI 0.46-1.48, p=0.37 and RR 0.79, 95% CI 0.43-1.48, p=0.33 respectively) (Supplementary Fig. 6d & e). Recent surgery was found to be associated with reduced risk of death with borderline significance (RR 0.79, 95% CI 0.63-0.99, p=0.047) from a meta-analysis of three studies (Supplementary Fig. 6f).

### Publication bias

The funnel plot on the effects of co-morbidities appeared slightly asymmetrical, but no significant publication bias was detected based on Eggers’ test (p-value=0.79). Similarly, no apparent asymmetry was visualised in the funnel plots on the effects of recent chemotherapy or surgery (Supplementary Fig. 7a-c).

## Discussion

This review and meta-analysis summarises the current data on patient characteristics and outcomes in cancer patients with COVID-19. To our knowledge, it includes the largest number of cancer patients and is the first to extract count data from all included studies. We also excluded studies that focused on specific cancer types in order to conduct an unbiased analysis of the risk of death associated with particular cancer diagnoses. In the case of studies that had recruited patients from the same geographical locations with overlapping authors’ institutions, we took a conservative approach of only including one of these studies. Whilst we may have excluded more patients than necessary, we have ruled out including patients twice, which strengthens the quality of the research evidence generated. All of the large registry-based studies published to date, UKCCMP, CCC19, OnCovid, and CACOVID-19 are included [10][12][16][17]. None of the meta-analyses presented had 80% power to detect a RR of 1.2 but the analyses of age, sex, metastatic cancer, smoking history, all cancer types, all comorbidities apart from hypertension, and all cancer therapies apart from recent surgery had 80% power to detect a RR of 1.5. All analyses had 80% power to detect a RR of 2.

Consistent with previous meta-analyses of cancer patients by Liu and Zhang et al, we identified that age, male sex and the presence of co-morbidities were associated with an increased risk of death, although the meta-analysis of sex did not reach statistical significance [18][19]. Large population studies of non-cancer COVID-19 patients have also found male patients, increasing age and chronic co-morbidities to be risk factors for increased hospital mortality [4][20]. Our results also suggest ECOG PS of 3 or greater and presence of a smoking history as risk factors for increased death but these results were not significant. Assessment of ECOG PS is also poorly reproduced in retrospective studies and confounded by the rapid deterioration from acute SARS-CoV-2 infection in hospitalised patients.

Co-existent cardiovascular disease, chronic lung disease, hypertension and diabetes were all significantly associated with an increased risk of death in our analysis. Significant between-study heterogeneity observed in the cardiovascular disease and hypertension meta-analyses were largely driven by the study by de Azambuja. This could partly be explained by the higher proportion of patients with cardiovascular disease (44%) than the other studies (6%-21%) [15]. Metastatic cancer patients with COVID-19 also appeared be at a higher risk of death. However, this analysis did not meet statistical significance and could be driven by confounding bias given that patients with higher cancer stages tend to have worse prognoses and not all studies in the meta-analysis reported on COVID-19 related deaths only.

Our meta-analyses also supported previous reports that patients with haematological cancers are at higher risk compared to patients with solid cancers [18][21]. A high risk of death (34%) was also reported in a systematic review of over 3,000 haematological cancer patients [22]. Interestingly compared to the meta-analysis by Liu et al, our results were not statistically significant, despite showing a trend for increased risk. When compared to Liu’s study, our analysis included more patients (396 vs 656 haematological and 1,726 vs 2,896 solid cancer patients) and had fewer potentially overlapping studies recruiting from the same geographical areas. Both analyses are not adjusted for other risk factors such as age, sex, performance status and co-morbidities. These may be important in estimating the true impact of haematological cancers on risk of death following SARS-CoV-2 infection. Our analysis does suggest that the impact of haematological cancer diagnosis may have been over-estimated by the smaller earlier studies whilst acknowledging the significant heterogeneity present (*I*^2^ 70%, p=0.001). Breast, gastrointestinal and lung cancers were common in the included studies, but we found no evidence of increased mortality risk within these cancer types. We did however find a high proportion of death among thoracic cancer patients from the included studies (37.7%), which supported the findings from a multi-centre study of 200 thoracic cancer patients that reported a high mortality rate of 33% in their cohort [23].

A key question during this pandemic has been whether to delay or avoid cancer treatment during escalating viral transmission. There have been two previous meta-analyses that have focused on the effect of active or recent cancer treatment on outcomes [24][25]. Yekedüz et al found no evidence of an association between recent anti-cancer treatment and increased risk of death when summary statistics from univariate models were analysed. However, they did find a statistically significant association between recent chemotherapy and an increased risk of death in multivariate models [25]. Park et al also found evidence of an association between recent chemotherapy and an increased risk of death when analysing summary statistics from univariate and multivariate models [24]. The multivariate analysis by Yekedüz only included 1,398 patients and involved only a subset of 227 haematological cancer patients from Lee et al [25]. In comparison, our meta-analysis of the impact of recent chemotherapy involved 4,256 patients and included all of the published large registry-based studies [9][12][16][17]. Only one study from our meta-analysis found a borderline significant association between recent chemotherapy and an increased risk of death in RT-PCR positive patients [16]. Our larger analysis provided no evidence that recent chemotherapy is associated with an increased risk of death and was powered at 71% to detect an effect size of 1.2. Previous analyses including fewer patients will also have been underpowered. Whilst we can rule out large risks associated with recent anti-cancer treatment, generally there is a need for new and ongoing cohort studies registering COVID-19 positive cancer patients to report on outcomes in patients on and off active treatment in the 60 days prior to a positive COVID-19 diagnosis in order to provide a definitive answer on the impact of recent treatment and to examine risk in particular subgroups such as haematological cancer patients.

Whilst the analyses of other cancer therapies included fewer patients, recent radiotherapy, immunotherapy, targeted and hormonal therapy were not associated with an increased risk of death. Although the analysis for recent surgery showed a borderline significant association with a lower risk of death, this analysis was underpowered to detect an effect size of 0.79 and potential confounders such as patients’ age, ECOG PS and type of surgery were not considered in the included studies. In addition, none of the studies provided details on median time between surgery and COVID-19 diagnosis. The COVIDSurg collaboration have published work to show that timing between surgery and a SARS-CoV-2 positive swab had a large impact on outcome and complications and that surgery in COVID-19 patients is associated with a high rate of pulmonary complications and deaths [26][27]. This collaborative project has also recently published another study showing that cancer patients treated in hospitals with COVID-19-free surgical pathways had lower rates of pulmonary complications and post-operative infection compared to patients treated in hospitals with no defined pathway [28].

## Limitations

Our analyses had several limitations. We were only able to include a limited number of studies in the meta-analyses for age, ECOG PS, smoking status, recent surgery, radiotherapy and hormonal therapy due to the lack of summary statistics from other studies for these variables. Statistical power to detect moderate effects (RRs) between 1.2 and 1.5 was low for the analyses of the impact of the different anti-cancer therapies on mortality. We were also unable to conduct further meta-analyses of odds ratios as most of the studies had adjusted their reported results for different variables. In addition, our review did not cover the rates and outcomes of haematological cancer patients who received either bone marrow or stem cell transplant or immunomodulatory medication due to the lack of data available for these therapies. Therefore, there is a need for further studies to evaluate the risks of patients receiving these therapies.

## Conclusion

Our meta-analysis of up to 5,678 patients has identified older age and presence of co-morbidities including cardiovascular, chronic lung conditions, diabetes and hypertension as significant risk factors among cancer patients with COVID-19. There was no clear increase in risk of death for a particular cancer type, although there was a trend towards haematological cancers being at higher risk compared to solid cancers. There was also no increase in risk of death with any recent anti-cancer therapy, including chemotherapy.

## Data Availability

Data generated or analysed during this study are included in this published article and its supplementary information files

## Notes

### Competing Interest Statement

DJP received lecture fees from ViiV Healthcare, Roche, Falk and Bayer Healthcare and travel expenses from BMS, MSD and Bayer Healthcare; consulting fees for Mina Therapeutics, EISAI, H3B, DaVolterra, Roche, and Astra Zeneca; received research funding (to institution) from MSD and BMS. LYWL has previously received speaker honorarium from the Merck group and Servier.

### Funding Statement

DJP is supported by grant funding from the Wellcome Trust Strategic Fund (PS3416) and acknowledges infrastructural support by the Cancer Research UK Imperial Centre and the Imperial NIHR BRC. CP and HC acknowledge funding from Bowel Cancer UK, ISC acknowledges funding from the Birmingham Cancer Research UK Centre.

## References

[1] Lake MA. What we know so far: COVID-19 current clinical knowledge and research. Clin Med (Northfield Il) 2020;20:124 LP–127. https://doi.org/10.7861/clinmed.2019-coron.

[2] Pentheroudakis G, Jordan K, Lordick F, Douillard JY, Peters S. What should medical oncologists know about COVID-19. Eur Soc Med Oncol, https://www.esmo.org/newsroom/covid-19-and-cancer/q-a-on-covid-19; 2020 [accessed March 31, 2020].

[3] World Health Organisation. COVID-19 weekly epidemiological update, https://www.who.int/emergencies/diseases/novel-coronavirus-2019/situation-reports; 2020 [accessed December 11, 2020].

[4] Docherty AB, Harrison EM, Green CA, Hardwick HE, Pius R, Norman L, et al. Features of 20L133 UK patients in hospital with covid-19 using the ISARIC WHO Clinical Characterisation Protocol: prospective observational cohort study. BMJ 2020;369:m1985. https://doi.org/10.1136/bmj.m1985.

[5] Wise J. Covid-19: Cancer mortality could rise at least 20% because of pandemic, study finds. BMJ 2020;369:m1735. https://doi.org/10.1136/bmj.m1735.

[6] Cancer Research UK. Cancer Research UK Cancer Patient Experience Survey 2020- The impact of COVID-19 on cancer patients in the UK. 2020.

[7] Aapro Matti, Addeo A AP et al. Cancer Patient Management during the COVID-19 Pandemic. ESMO Guidel 2020, https://www.esmo.org/guidelines/cancer-patient-management-during-the-covid-19-pandemic; 2020 [accessed November 17, 2020].

[8] Moher D, Liberati A, Tetzlaff J, Altman DG, Group TP. Preferred Reporting Items for Systematic Reviews and Meta-Analyses: The PRISMA Statement. PLoS Med 2009;6:e1000097. https://doi.org/10.1371/journal.pmed.1000097.

[9] Lee LYW, Cazier JB, Starkey T, Turnbull CD, Kerr R, Middleton G. COVID-19 mortality in patients with cancer on chemotherapy or other anticancer treatments: a prospective cohort study. Lancet 2020. https://doi.org/10.1016/S0140-6736(20)31173-9.

[10] Lee LYW, Cazier J-B, Starkey T, Briggs SEW, Arnold R, Bisht V, et al. COVID-19 prevalence and mortality in patients with cancer and the effect of primary tumour subtype and patient demographics: a prospective cohort study. Lancet Oncol 2020;21:1309–16. https://doi.org/10.1016/S1470-2045(20)30442-3.

[11] Pinato DJ, Lee AJX, Biello F, Seguí E, Aguilar-Company J, Carbó A, et al. Presenting Features and Early Mortality from SARS-CoV-2 Infection in Cancer Patients during the Initial Stage of the COVID-19 Pandemic in Europe. Cancers (Basel) 2020;12:1841. https://doi.org/10.3390/cancers12071841.

[12] Pinato DJ, Zambelli A, Aguilar-Company J, Bower M, Sng CCT, Salazar R, et al. Clinical Portrait of the SARS-CoV-2 Epidemic in European Patients with Cancer. Cancer Discov 2020;10:1465 LP–1474. https://doi.org/10.1158/2159-8290.CD-20-0773.

[13] Balduzzi S, Rücker G, Schwarzer G. How to perform a meta-analysis with R: a practical tutorial. Evid Based Ment Heal 2019;22:153 LP–160. https://doi.org/10.1136/ebmental-2019-300117.

[14] Viechtbauer W. Conducting Meta-Analyses in R with the metafor Package. J Stat Software; Vol 1, Issue 3 2010. https://doi.org/10.18637/jss.v036.i03.

[15] de Azambuja E, Brandão M, Wildiers H, Laenen A, Aspeslagh S, Fontaine C, et al. Impact of solid cancer on in-hospital mortality overall and among different subgroups of patients with COVID-19: a nationwide, population-based analysis. ESMO Open 2020;5:e000947. https://doi.org/10.1136/esmoopen-2020-000947.

[16] Lièvre A, Turpin A, Ray-Coquard I, Le Malicot K, Thariat J, Ahle G, et al. Risk factors for Coronavirus Disease 2019 (COVID-19) severity and mortality among solid cancer patients and impact of the disease on anticancer treatment: A French nationwide cohort study (GCO-002 CACOVID-19). Eur J Cancer 2020;141:62–81. https://doi.org/10.1016/j.ejca.2020.09.035.

[17] Kuderer NM, Choueiri TK, Shah DP, Shyr Y, Rubinstein SM, Rivera DR, et al. Clinical impact of COVID-19 on patients with cancer (CCC19): a cohort study. Lancet 2020. https://doi.org/10.1016/S0140-6736(20)31187-9.

[18] Liu Y, Lu H, Wang W, Liu Q, Zhu C. Clinical risk factors for mortality in patients with cancer and COVID-19: a systematic review and meta-analysis of recent observational studies. Expert Rev Anticancer Ther 2020:1–13. https://doi.org/10.1080/14737140.2021.1837628.

[19] Zhang H, Han H, He T, Labbe KE, Hernandez A V, Chen H, et al. Clinical Characteristics and Outcomes of COVID-19-Infected Cancer Patients: A Systematic Review and Meta-Analysis. JNCI J Natl Cancer Inst 2020. https://doi.org/10.1093/jnci/djaa168.

[20] Deng G, Yin M, Chen X, Zeng F. Clinical determinants for fatality of 44,672 patients with COVID-19. Crit Care 2020;24:179. https://doi.org/10.1186/s13054-020-02902-w.

[21] Venkatesulu BP, Thoguluva Chandrasekar V, Giridhar P, Advani P, Sharma A, Hsieh CE, et al. A systematic review and meta-analysis of cancer patients affected by a novel coronavirus. MedRxiv 2020:2020.05.27.20115303. https://doi.org/10.1101/2020.05.27.20115303.

[22] Vijenthira A, Gong IY, Fox TA, Booth S, Cook G, Fattizzo B, et al. Outcomes of patients with hematologic malignancies and COVID-19: A systematic review and meta-analysis of 3377 patients. Blood 2020. https://doi.org/10.1182/blood.2020008824.

[23] Garassino MC, Whisenant JG, Huang L-C, Trama A, Torri V, Agustoni F, et al. COVID-19 in patients with thoracic malignancies (TERAVOLT): first results of an international, registry-based, cohort study. Lancet Oncol 2020;21:914–22. https://doi.org/10.1016/S1470-2045(20)30314-4.

[24] Park R, Lee SA, Kim SY, de Melo AC, Kasi A. Association of active oncologic treatment and risk of death in cancer patients with COVID-19: a systematic review and meta-analysis of patient data. Acta Oncol (Madr) 2020:1–7. https://doi.org/10.1080/0284186X.2020.1837946.

[25] Yekedüz E, Utkan G, Ürün Y. A systematic review and meta-analysis: the effect of active cancer treatment on severity of COVID-19. Eur J Cancer 2020;141:92–104. https://doi.org/10.1016/j.ejca.2020.09.028.

[26] Collaborative Covids. Delaying surgery for patients with a previous SARS-CoV-2 infection. BJS (British J Surgery) 2020;107:e601–2. https://doi.org/10.1002/bjs.12050.

[27] Nepogodiev D, Bhangu A, Glasbey JC, Li E, Omar OM, Simoes JFF, et al. Mortality and pulmonary complications in patients undergoing surgery with perioperative SARS-CoV-2 infection: an international cohort study. Lancet 2020;396:27–38. https://doi.org/10.1016/S0140-6736(20)31182-X.

[28] Glasbey JC, Bhangu A. Elective Cancer Surgery in COVID-19–Free Surgical Pathways During the SARS-CoV-2 Pandemic: An International, Multicenter, Comparative Cohort Study. J Clin Oncol 2020:JCO.20.01933. https://doi.org/10.1200/JCO.20.01933.

[29] Mehta A, Vasudevan S, Parkash A, Sharma A, Vashist T, Krishna V. COVID-19 Mortality in Cancer Patients: A Report from a Tertiary Cancer Centre in India. MedRxiv 2020:2020.09.14.20194092. https://doi.org/10.1101/2020.09.14.20194092.

[30] Stroppa EM, Toscani I, Citterio C, Anselmi E, Zaffignani E, Codeluppi M, et al. Coronavirus disease-2019 in cancer patients. A report of the first 25 cancer patients in a western country (Italy). Future Oncol 2020;16:1425–32. https://doi.org/10.2217/fon-2020-0369.

[31] de Joode K, Dumoulin DW, Tol J, Westgeest HM, Beerepoot L V, van den Berkmortel FWPJ, et al. Dutch Oncology COVID-19 consortium: Outcome of COVID-19 in patients with cancer in a nationwide cohort study. Eur J Cancer 2020;141:171–84. https://doi.org/10.1016/j.ejca.2020.09.027.

[32] Ramachandran P, Kathirvelu B, Chakraborti A, Gajendran M, Zhahid U, Ghanta S, et al. COVID-19 in Cancer Patients From New York City: A Comparative Single Center Retrospective Analysis. Cancer Control 2020;27:1073274820960457. https://doi.org/10.1177/1073274820960457.

[33] Russell B, Moss C, Papa S, Irshad S, Ross P, Spicer J, et al. Factors Affecting COVID-19 Outcomes in Cancer Patients: A First Report From Guy’s Cancer Center in London. Front Oncol 2020;10:1279. https://doi.org/10.3389/fonc.2020.01279.

[34] de Melo AC, Thuler LCS, da Silva JL, de Albuquerque LZ, Pecego AC, Rodrigues L de OR, et al. Cancer inpatients with COVID-19: A report from the Brazilian National Cancer Institute. PLoS One 2020;15:e0241261. https://doi.org/10.1371/journal.pone.0241261.

[35] Wang J, Song Q, Chen Y, Wang Z, Chu Q, Gong H, et al. Systematic investigations of COVID-19 in 283 cancer patients. MedRxiv 2020:2020.04.28.20083246. https://doi.org/10.1101/2020.04.28.20083246.

[36] Pinto C, Berselli A, Mangone L, Damato A, Iachetta F, Foracchia M, et al. SARS-CoV- 2 Positive Hospitalized Cancer Patients during the Italian Outbreak: The Cohort Study in Reggio Emilia. Biology (Basel) 2020;9:181. https://doi.org/10.3390/biology9080181.

