## Supplementary Information for "Influence of clinical characteristics and anti-cancer therapy on outcomes from SARS-CoV-2 infection: a systematic review and meta-analysis of 5,678 cancer patients"

**Supplementary Table 1. PRISMA Checklist**

**Supplementary Table 2. Characteristics of cancer patients with COVID-19.**

**Supplementary Table 3. Results from the meta-analysis of the impact of COVID-19 cancer patients with gastrointestinal, breast and thoracic cancer on the risk of death (excluding the study by Lièvre which analysed solid organ cancers only). Abbreviations**: CI- confidence interval

**Supplementary Fig. 1. Forest plot of the pooled proportion of male cancer patients with COVID-19.** Abbreviations: CI- Confidence interval

**Supplementary Fig 2. Forest plot of contribution of male sex to risk of death in COVID-19 cancer patients.** Abbreviations: CI- Confidence interval, RR- Risk ratio

**Supplementary Fig 3. Forest plot of the contribution of ECOG PS to risk of death in COVID-19 cancer patients when comparing those scoring 3 or higher to those scoring less than 3.** Abbreviations: CI- Confidence interval, ECOG- Eastern Cooperative Oncology Group, PS- performance status, RR- risk ratio

**Supplementary Fig. 4. Forest plot of the contribution of smoking to risk of death in COVID-19 cancer patients who have a smoking history versus no smoking history.** Abbreviations: CI- Confidence interval, RR- Risk ratio

**Supplementary Fig. 5. Forest plot of the contribution of metastatic disease to risk of death in COVID-19 cancer patients.** Abbreviations: CI- Confidence interval, RR- Risk ratio

**Supplementary Fig. 6. Forest plot of risk of death in COVID-19 cancer patients who had recent anti-cancer therapy. a) Contribution of any recent anti-cancer therapy. Recent was defined as ongoing treatment at time of COVID-19 diagnosis or within 60 days of their diagnosis. b-f) Contribution of recent immunotherapy, targeted therapy, radiotherapy, hormonal therapy and surgery.** Abbreviations: CI- Confidence interval, RR- Risk ratio

**Supplementary Fig. 7. Funnel plot to assess publication bias on the effects of a) presence of one or more co-morbidities b) recent chemotherapy c) recent surgery on the risk of death in COVID-19 cancer patients.** Abbreviations: RR- Risk ratio

| **Section** | | **No.** | **Checklist item** | | | **Reported in page no.** |
| --- | --- | --- | --- | --- | --- | --- |
| **TITLE** | | | | | |  |
| Title | | 1 | Identify the report as a systematic review, meta-analysis, or both. | | | 1 |
| **ABSTRACT** | | | | | |  |
| Structured summary | | 2 | Provide a structured summary including, as applicable: background; objectives; data sources; study eligibility criteria, participants, and interventions; study appraisal and synthesis methods; results; limitations; conclusions and implications of key findings; systematic review registration number. | | | 1 |
| **INTRODUCTION** | | | | | |  |
| Rationale | | 3 | Describe the rationale for the review in the context of what is already known. | | | 2 |
| Objectives | | 4 | Provide an explicit statement of questions being addressed with reference to participants, interventions, comparisons, outcomes, and study design (PICOS). | | | 2 |
| **METHODS** | | | | | |  |
| Eligibility criteria | | 5 | Specify study characteristics (e.g., PICOS, length of follow-up) and report characteristics (e.g., years considered, language, publication status) used as criteria for eligibility, giving rationale. | | | 2 |
| Information sources | | 6 | Describe all information sources (e.g., databases with dates of coverage, contact with study authors to identify additional studies) in the search and date last searched. | | | 2 |
| Search | | 7 | Present full electronic search strategy for at least one database, including any limits used, such that it could be repeated. | | | 2 |
| Study selection | | 8 | State the process for selecting studies (i.e., screening, eligibility, included in systematic review, and, if applicable, included in the meta-analysis). | | | 2-3 |
| Data collection process | | 9 | Describe method of data extraction from reports (e.g., piloted forms, independently, in duplicate) and any processes for obtaining and confirming data from investigators. | | | 3 |
| Data items | | 10 | List and define all variables for which data were sought (e.g., PICOS, funding sources) and any assumptions and simplifications made. | | | 3 |
| Summary measures | | 11 | State the principal summary measures (e.g., risk ratio, difference in means). | | | 3 |
| Synthesis of results | | 12 | Describe the methods of handling data and combining results of studies, if done, including measures of consistency (e.g., I^2^) for each meta-analysis. | | | 3 |
| Section | | No. | Checklist item | | | Reported in page no. |
| Risk of bias across studies | | 13 | Specify any assessment of risk of bias that may affect the cumulative evidence (e.g., publication bias, selective reporting within studies). | | | 3 |
| **RESULTS** | | | | |  | |
| Study selection | 14 | | | Give numbers of studies screened, assessed for eligibility, and included in the review, with reasons for exclusions at each stage, ideally with a flow diagram. | 4, Figure 1 | |
| Study characteristics | 15 | | | For each study, present characteristics for which data were extracted (e.g., study size, PICOS, follow-up period) and provide the citations. | 4, Table 1 | |
| Results of individual studies | 16 | | | For all outcomes considered (benefits or harms), present, for each study: (a) simple summary data for each intervention group (b) effect estimates and confidence intervals, ideally with a forest plot. | 4-7, Figures 2-5, Suppl. information | |
| Synthesis of results | 17 | | | Present results of each meta-analysis done, including confidence intervals and measures of consistency. | 4-7, Figures 2-5, Suppl. Information | |
| Risk of bias across studies | 18 | | | Present results of any assessment of risk of bias across studies (see Item 15). | 7, Suppl. information | |
| **DISCUSSION** | | | | |  | |
| Summary of evidence | 19 | | | Summarize the main findings including the strength of evidence for each main outcome; consider their relevance to key groups (e.g., healthcare providers, users, and policy makers). | 7-10 | |
| Limitations | 20 | | | Discuss limitations at study and outcome level (e.g., risk of bias), and at review-level (e.g., incomplete retrieval of identified research, reporting bias). | 10 | |
| Conclusions | 21 | | | Provide a general interpretation of the results in the context of other evidence, and implications for future research. | 10 | |
| **FUNDING** | | | | |  | |
| Funding | 22 | | | Describe sources of funding for the systematic review and other support (e.g., supply of data); role of funders for the systematic review. | 3 | |

**Supplementary Table 1. PRISMA Checklist**

| **Variable\Study** | **Lee** | **^a^Lee** | **Kuderer** | **de**  **Azambuja** | **Mehta** | **Stroppa** | **Pinato** | **^a^Pinato** | **de Joode** | **Ramachandran** | **Russell** | **de Melo** | **Wang** | **Lièvre** | **Pinto** |
| --- | --- | --- | --- | --- | --- | --- | --- | --- | --- | --- | --- | --- | --- | --- | --- |
| Smoking status of COVID-19 cancer patients (n, % ever smokers) | NS | 311, 29.8% | 369, 39.8% | 100, 8.4%  (current-smoker) | NS | 13, 52%  (current-smoker) | 112, 54.9% | 380, 42.7% | 179, 51% | 22, 41.5%  (current-smoker) | 50, 32.1% | 40, 22.1% | 65, 23% | 564, 43.8% | 52, 37.7% |
| ECOG performance status of cancer patients (n,%) | NS | NS | -PS 0-1 (614, 66%)  -PS 2  (72, 8%)  -PS 3-4 (46, 5%) | NS | NS | NS | NS | NS | NS | NS | -PS 0-1 (62, 40%)  -PS 2 (33, 21%)  -PS 3-4 (20, 13%) | NS | NS | -PS 0-1 (547, 42%)  -PS 2 (247, 19%)  -PS 3-4 (137, 11%) | NS |
| No. of patients with comorbidities (n,%)  CVD  Chronic lung disease  Diabetes  Hypertension | 631, 79%  104, 13%  61, 8%  131, 16%  247, 31% | 664, 64%  145, 14%  80, 8%  178, 17%  343, 33% | 742, 80%  NS  NS  NS  NS | 739, 83%^b^  395, 44%^b^  204, 23%^b^  208, 23%^b^  386, 43%^b^ | 86, 46%  11, 6%  2, 1%  34, 18%  45, 24% | 19, 76%  NS  7, 28%  8, 32%  16, 64% | 161, 78%  44, 22%  34, 17%  46, 23%  88, 43% | 670, 75%  190, 21%  119, 13%  181, 20%  386, 43% | NS  190, 54%  46, 13%  55, 16%  NS | NS  10, 19%  5, 9%  NS  44, 83% | 113, 72%  29, 19%  25, 16%  35, 22%  74, 47% | 72, 40%  NS  7, 4%  31, 17%  77, 43% | NS  32, 11%  20, 7%  39, 14%  94, 33% | 1114, 86%  194, 15%  124, 10%  241, 19%  529, 41% | NS  77, 56%  18, 13%  37, 27%  105, 76% |
| No. of cancer patients with metastatic/advanced cancer (n, %) | 347, 43.4% | NS | NS | NS | 50, 26.9% | 19, 76%^c^ | 82, 40.2% | 351, 39.4% | 112, 31.9% | 13, 24.5% | 63, 40.4% | 90, 49.7% | 101, 35.7%^c^ | 758, 58.8% | 11/14, 78.6%^d^ |
| No. of patients with solid organ cancers (n, %)  Gastrointestinal  Thoracic  Breast | 631, 79%  150, 19%  90, 11%  102, 13% | 817, 78%  219, 21%  122, 12%  143, 14% | 654, 71%^e^  108, 12%  91, 10%  191, 21% | 892, 100%^b^  NS  NS  NS | 153, 82%  40, 22%  17, 9%  19, 10% | 22, 88%  6, 24%  8, 32%  2, 8% | 185, 91%  47, 23%  36, 18%  27, 13% | 753, 85%  190, 21%  119, 13%  162, 18% | 238, 68%  41, 12%  51, 15%  47, 13% | 41, 77%  8, 15%  7, 13%  10, 19% | 128, 82%  21, 14%  17, 11%  24, 15% | 138, 76%  24, 13%  7, 4%  40, 22% | NS  74, 26%  51, 18%  38, 13% | 1289, 100%  470, 37%  311, 24%^f^  173, 13% | NS  29, 21%  9, 7%  27, 20% |
| No. of patients with haematological cancers (n, %) | 169, 21% | 227, 22% | 167, 18%^e^ | 30, 3% | 33, 18% | 2, 8% | 19, 10% | 137, 15% | 113, 32% | 11, 21% | 28, 18% | 34, 19% | 21, 7% | 0, 0% | NS |
| No. of patients who had recent cancer therapy (n,%)  Chemotherapy  Immunotherapy  Radiotherapy  Targeted therapy  Hormonal therapy  Surgery  Definition of recent therapy | 518, 65%  281, 35%  44, 6%  76, 10%  72, 9%  64, 8%  29, 4%  Within 4 weeks of infection | NS  349, 33%  39, 4%  86, 8%  91, 9%  NS  36, 3%  Within 4 weeks of infection | 366, 39%  160, 17%  38, 4%  12, 1%  75, 8%  85, 9%  2, 0.2%  Within 4 weeks of infection | NS  NS  NS  NS  NS  NS  NS  NS | 112, 60%  69, 37%  11, 6%  21, 11%  6, 3%  NS  31, 17%  Within 1 month of infection | 12, 48%  8, 32%  4, 16%  NS  NS  NS  NS  At time of infection | 102, 50%  38, 19%  5, 3%  65, 32%  13, 6%  NS  96, 47%  At time of infection | 479, 54%  206, 23%  56, 6%  33, 4%  93, 10%  92, 10%  NS  Within 4 weeks of infection | 165, 47%  NS  NS  NS  NS  NS  NS  Within 30 days of infection | 21, 40%  11, 21%  2, 4%  1, 2%  1,2%  6, 11%  1, 2%  NS | 81, 52%  45, 29%  7, 5%  NS  18, 12%  NS  NS  NS | NS  63, 35%  9, 5%  10, 6%  9, 5%  20, 11%  12, 7%  Within last 60 days of hospitalisation | 95, 34%  46, 16%  2, 0.7%  NS  12, 4%  NS  23, 8%  Within 3 months of hospitalisation | 491, 38%  385, 30%  62, 5%  95, 7%  114, 9%  27, 2%  56, 4%  Within 4 weeks of infection | 14, 10%  12, 9%  1, 0.7%  NS  NS  2, 1%  NS  Within 60 days of hospitalisation |

**Supplementary Table 2. Characteristics of cancer patients with COVID-19.** Types of co-morbidities, cancer types and anti-cancer therapy shown in this table reflect those included in the meta-analyses only. **Abbreviations:** CVD- cardiovascular disease, ECOG- Eastern Cooperative Oncology Group, NS- not specified**,** PS- performance status

^a^ Second publication from the same registry study

^b^ Numbers shown here refer to cases in hospital only

^c^ Includes locally advanced and advanced cases

^d^ Numbers shown here are for patients on active treatment only

^e^ Numbers shown here are cases with solid organ or haematological cancers only and does not include cases with multiple cancers

^f^ Includes mesothelioma cases and others (n=52)

|  | **Risk Ratio (RR)** | **95% CI** | **p-value** |
| --- | --- | --- | --- |
| **Gastrointestinal Cancer** | 1.09 | 0.64-1.84 | 0.69 |
| **Breast Cancer** | 0.82 | 0.39-1.76 | 0.55 |
| **Thoracic Cancer** | 1.10 | 0.63-1.90 | 0.69 |

**Supplementary Table 3. Results from the meta-analysis of the impact of COVID-19 cancer patients with gastrointestinal, breast and thoracic cancer on the risk of death (excluding the study by Lièvre which analysed solid organ cancers only). Abbreviations**: CI- confidence interval

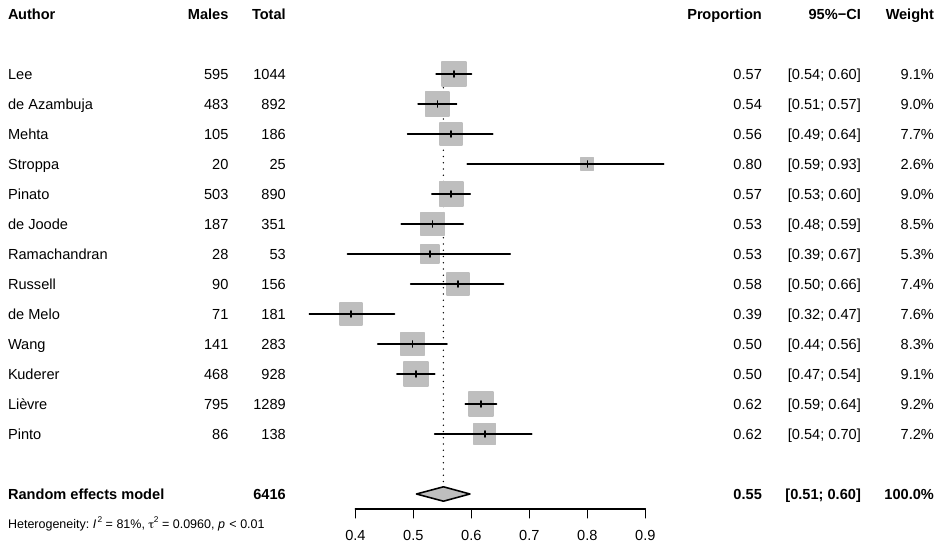

**Supplementary Fig. 1. Forest plot of the pooled proportion of male cancer patients with COVID-19**

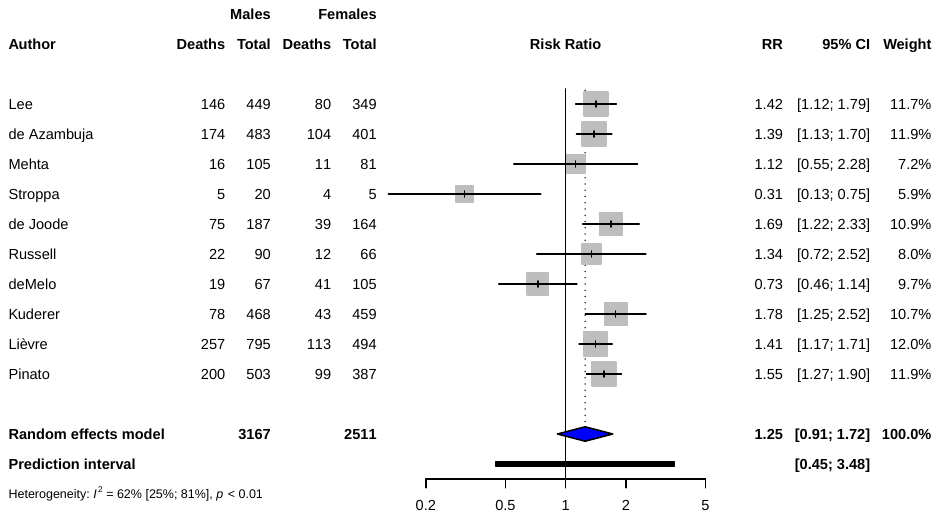
**Supplementary Fig 2. Forest plot of contribution of male sex to risk of death in COVID-19 cancer patients**

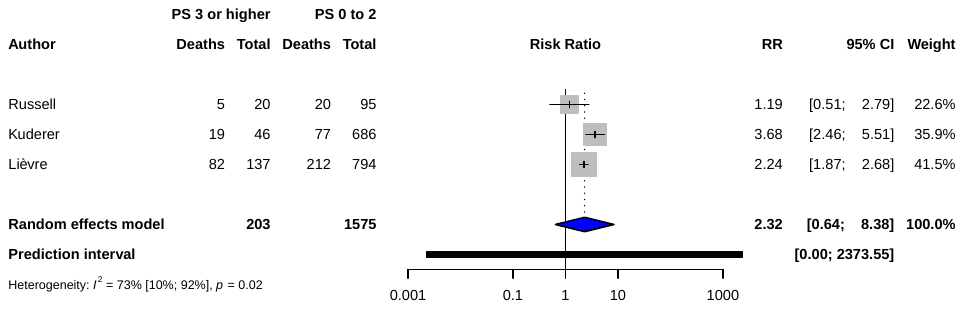

**Supplementary Fig 3. Forest plot of the contribution of ECOG PS to risk of death in COVID-19 cancer patients when comparing those scoring 3 or higher to those scoring less than 3. Abbreviations: ECOG-** Eastern Cooperative Oncology Group**, PS-** performance status

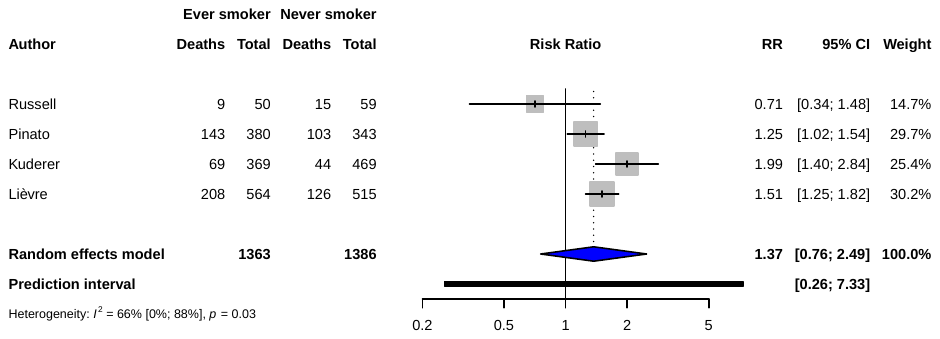
 **Supplementary Fig. 4. Forest plot of the contribution of smoking to risk of death in COVID-19 cancer patients who have a smoking history versus no smoking history**

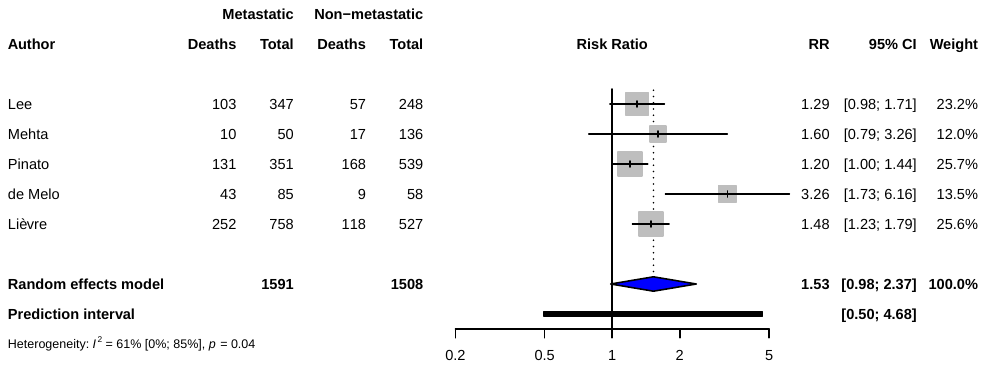
 **Supplementary Fig. 5. Forest plot of the contribution of metastatic disease to risk of death in COVID-19 cancer patients**

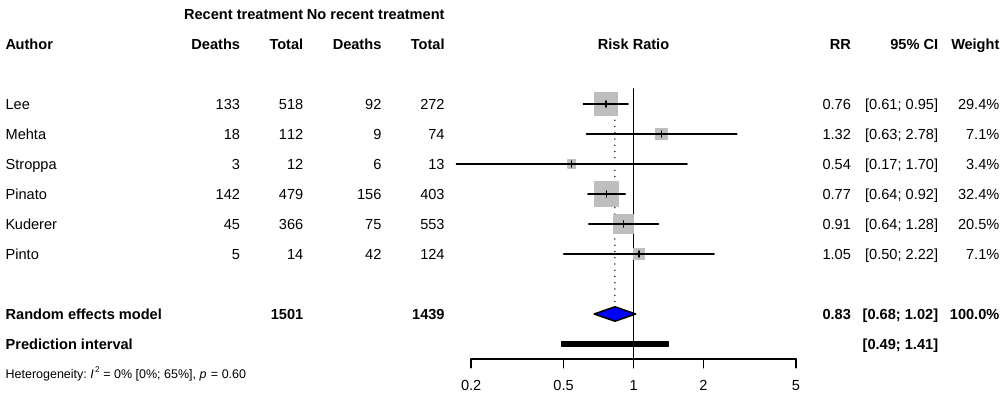

6b)
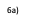

6a)

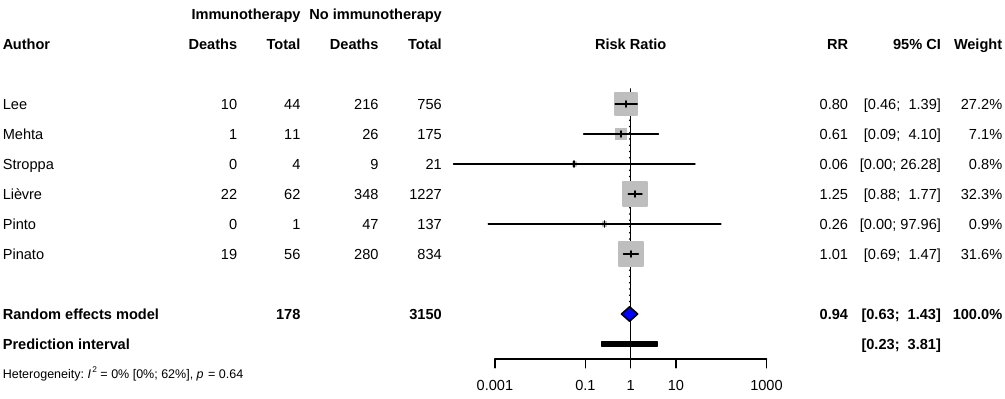

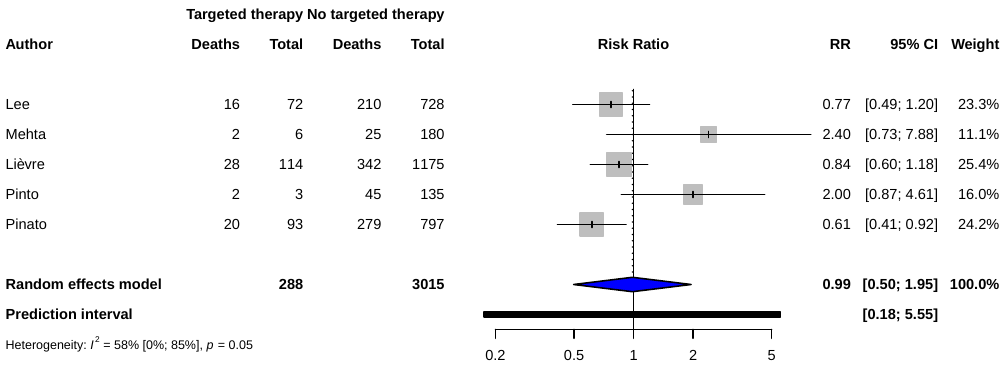

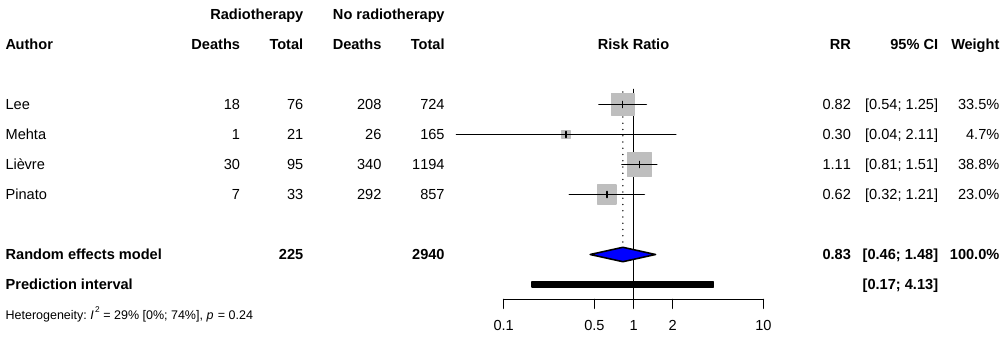

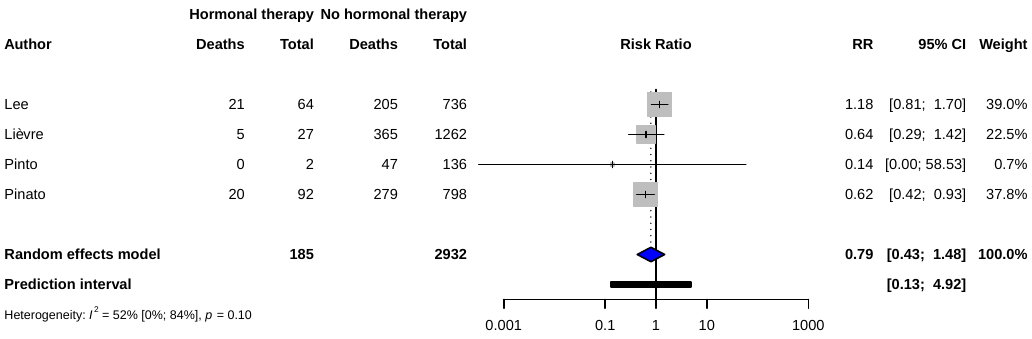

6c)
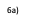

6e)

6d)

6f)

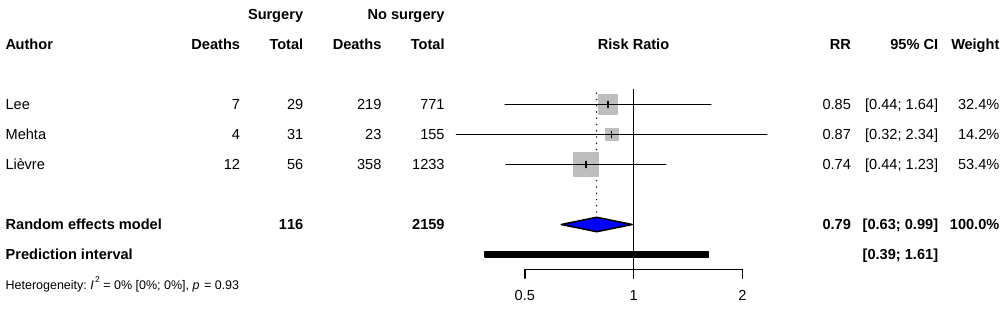

**Supplementary Fig. 6. Forest plot of risk of death in COVID-19 cancer patients who had recent anti-cancer therapy. a) Contribution of any recent anti-cancer therapy. Recent was defined as ongoing treatment at time of COVID-19 diagnosis or within 60 days of their diagnosis. b-f) Contribution of recent immunotherapy, targeted therapy, radiotherapy, hormonal therapy and surgery**

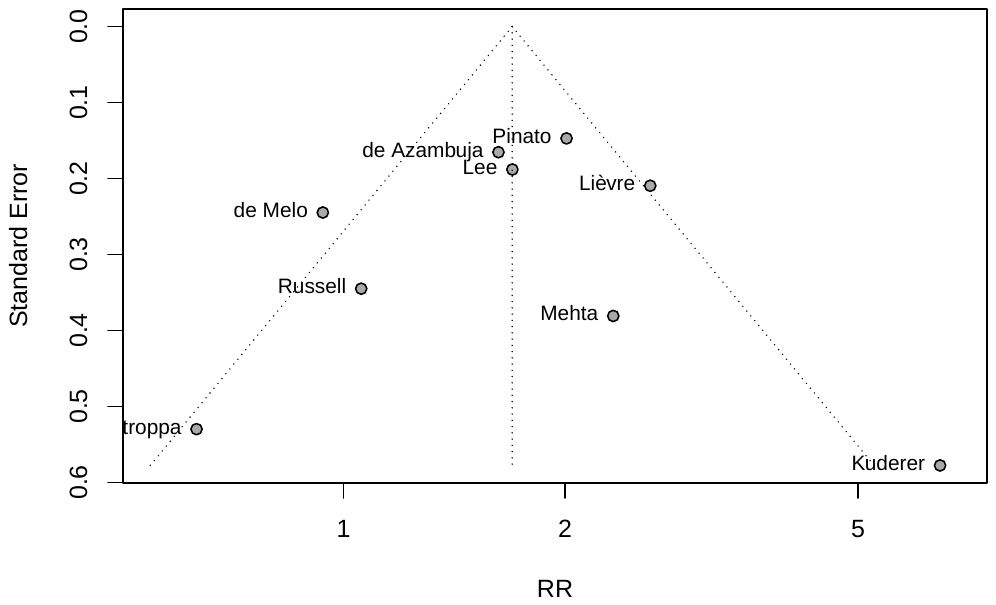

7a)

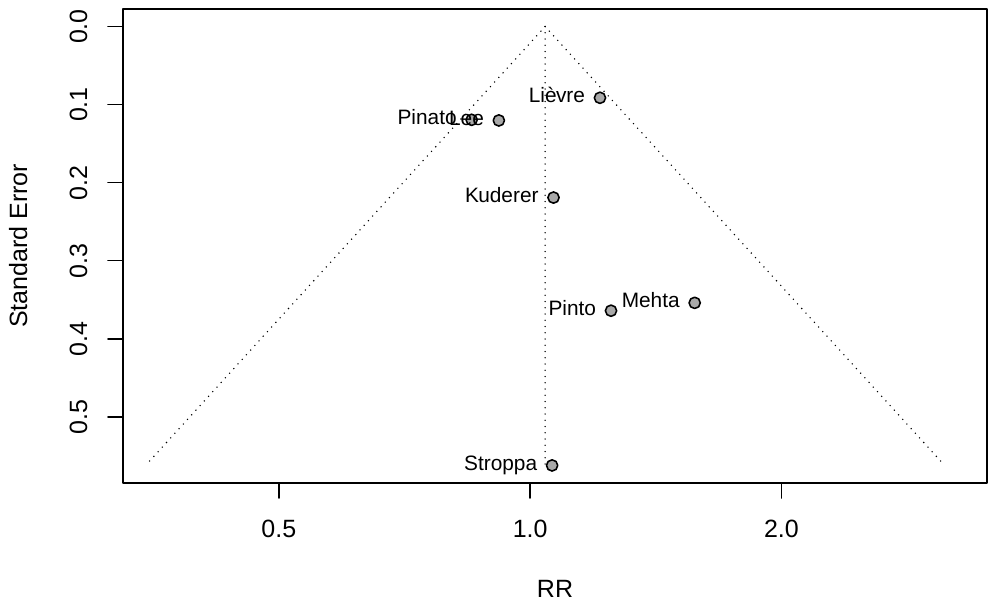

7b)
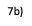

7b)
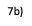

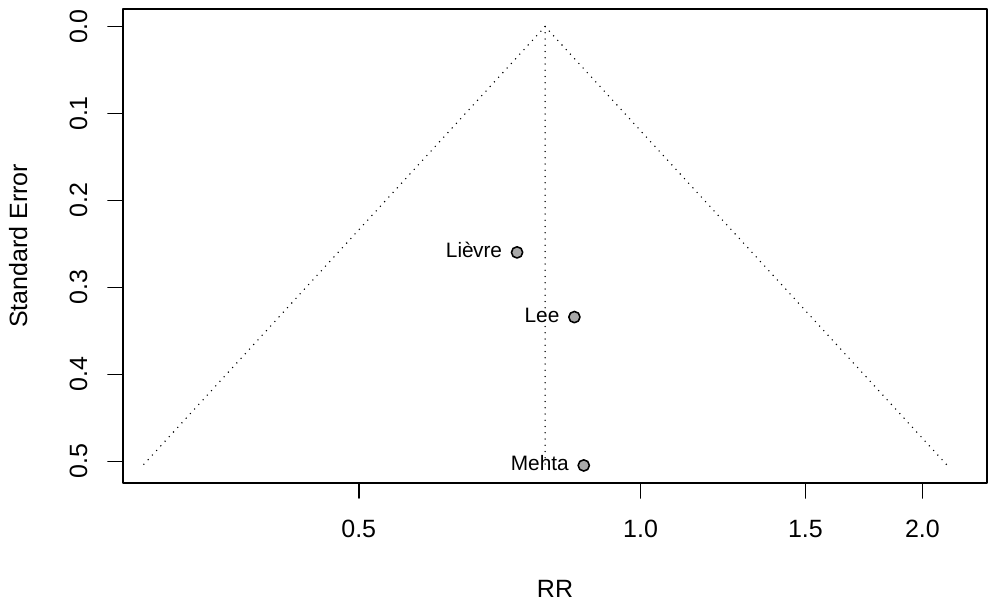

7c)
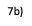

**Supplementary Fig. 7. Funnel plot to assess publication bias on the effects of a) presence of one or more co-morbidities b) recent chemotherapy c) recent surgery on the risk of death in COVID-19 cancer patient**
